# Pandemic Influenza Protection Before Day One: Modeling the Impact of a Broadly Cross-Reactive Seasonal Vaccine

**DOI:** 10.64898/2026.07.22.26358670

**Authors:** Luis Mier-y-Teran-Romero, David P. Durham, John Treanor, Nicolas J. Swanson, Lakshmi Jayashankar, Samson Lee

## Abstract

Pandemic influenza poses a constant threat to domestic and global public health with the potential of high morbidity and mortality. Pandemic virus-matched vaccines, though potentially highly efficacious, require a lead time before they can be mass-produced and distributed to the population. For this reason, mitigation strategies that provide protection ahead of widespread viral transmission are highly desirable. Here, we model the impact of a broadly cross-reactive seasonal influenza vaccine on pandemic influenza spread in the U.S. Our findings indicate that as the cross-protective efficacy of the seasonal vaccine against the pandemic variant increases, total hospitalizations are concomitantly reduced. For example, we find that even at typical seasonal vaccine uptake and modest vaccine efficacies against infection and hospitalization with the pandemic variant, total hospitalizations can be decreased by 30% or more across the scenarios considered. In addition, the hospitalization risk for recipients is at least 35% lower than the risk experienced by unvaccinated persons. These results demonstrate that a broadly protective influenza vaccine could serve as a critical component of pandemic preparedness strategies.

## Introduction

Pandemic influenza represents a priority threat to U.S. health security [1]. Pandemics are caused by newly emerging influenza viruses–originating from zoonotic spillover or antigenic-shift events–that pose a threat to human populations due to the lack of population immunity [2]. In the years since 1918, the world has experienced five influenza pandemics, occurring in 1918, 1957, 1968, 1977 and 2009; each with a varying degree of morbidity and mortality [3, 4, 5, 6, 7, 8]. While it is impossible to accurately predict precisely when an emergent virus will give rise to a pandemic, antigenic changes and zoonotic spillover events occur constantly and make pandemic preparedness an urgent necessity [9]. Strengthening surveillance systems, increasing throughput of laboratory testing capabilities, and developing new vaccines and therapeutics remain critical priorities [1]. Even more so, to counter the pandemic potential of influenza viruses, it is necessary to combine mitigation strategies effectively and to consider novel approaches as new technologies arise.

The U.S. Department of Health and Human Services (HHS) 2017 Pandemic Influenza Plan for countering pandemic influenza includes a combination of community mitigation measures, matched vaccines, and therapeutics [1]. Pandemic strain-matched vaccines can be highly efficacious at limiting transmission in the population and improving clinical outcomes among recipients. However, strain-matched vaccines require several months of lead time before they can be distributed on a large scale [10, 11, 12, 13, 14]. Although the HHS 2017 Pandemic Influenza Plan foresees the capability to deliver finished vaccine doses by 12 weeks after the declaration of a pandemic, this timeline may be insufficient for a highly transmissible virus. In fact, there is a large body of evidence suggesting that a lower efficacy vaccine that is deployed sufficiently early can mitigate morbidity and mortality to a greater extent than a highly efficacious vaccine delivered on a delayed timeline [14, 15].

Seasonal, non-pandemic, influenza viruses also cause substantial morbidity and mortality in the US and worldwide. Their transmission is tempered by the presence of partial immunity in the population, generated by the circulation of related viruses in previous seasons. In the US, established vaccination infrastructure and public awareness support routine seasonal immunization. Therefore, a valuable addition to pandemic preparedness would be a seasonal vaccine with cross-reactivity that offers broad-protection against a multitude of virus variants, henceforth referred to as a “Before Day One Vaccine” [13]. Such a vaccine would help to build a reservoir of immunity among the population, potentially blunting and slowing the peak of an outbreak to allow time for manufacturing and distribution of a strain-matched vaccine.

To develop a Before Day One vaccine, possible mechanisms of action include targeting conserved regions of influenza viral proteins, developing multivalent vaccines encoding antigens from many subtypes and lineages, boosting cross-reactive T-cell responses, leveraging virus-like particle platforms to induce broad immunity [16, 17, 18, 19, 20, 21], among others. The goal is to achieve cross-reactivity and provide broad coverage against disparate seasonal variants, in addition to variants with pandemic potential that have yet to emerge. An additional benefit would be reducing the need for a complete vaccine reformulation each season, as well as reducing the risk of a seasonal strain-vaccine mismatch. Progress on broadly protective influenza vaccines is moving forward, with recent technological advances bringing candidate vaccines to animal and phase I clinical trials [16].

In this work, we model the impact of a broadly cross-reactive Before Day One vaccine on influenza pandemic spread in the US across three influenza pandemic scenarios. It is assumed that this vaccine is provided seasonally and thus shifts the immune landscape from fully naïve to one with partial protection at the time of pandemic virus emergence. We consider the additional impact that a strain-matched vaccine, with higher efficacy but delayed availability, would have on the incidence epidemic curve as well as on severe clinical outcomes. Our results demonstrate that even at modest Before Day One vaccine efficacies against infection and hospitalization, the total hospitalization burden can be reduced across the scenarios considered. We also demonstrate individual-level benefits through a considerable reduction in hospitalization rate among vaccine recipients, when compared to unvaccinated individuals. Our results show that broadly protective Before Day One vaccines have the potential to become a key tool in influenza pandemic preparedness.

## Methods

### Mathematical Model

We formulated a deterministic compartmental model to study influenza virus transmission. The model stratifies the 2024 U.S. population into five age categories (0-y4rs, 5-17yrs, 18-49yrs, 50-64yrs and 65+ yrs). The population is further stratified into disease compartments (susceptible, exposed, infected, and recovered) and into compartments indicating vaccination status (Supplement Fig. S1). We considered two categories of vaccination: Before Day One and strain-matched. We included the distribution of a Before Day One Vaccine with age-specific uptake set to typical levels seen in the U.S. over the 2021 – 2024 influenza seasons [22, 23]. In addition to Before Day One vaccination, we also considered situations in which a pandemic strain-matched vaccine is distributed in the population, incorporating a time delay in distribution to account for its manufacturing and distribution. We assumed that both vaccines provide some level of immunity against infection with the pandemic virus and mitigate the likelihood of hospitalization among breakthrough infections. As such, each vaccine is characterized by two efficacies: an efficacy against infection and a second one against hospitalization.

We focused on hospitalizations to compare the impact of different mitigation strategies. The number of hospitalizations is a key public health metric that serves as a proxy for nearly all clinical outcomes among infected individuals. Furthermore, reducing hospitalization incidence allows for the conservation of resources and capacity. To this end, we implemented age-specific pandemic infection hospitalization rates (IHR), drawing on estimates from previous seasonal and pandemic influenza as well as from COVID-19.

### Scenarios

We considered several pandemic scenarios in addition to different combinations of vaccine mitigations. First, we examined three pandemic scenarios that we label as ‘2009-like’, ‘1968-like’, and ‘1918-like’, arranged by increasing infectiousness. For each of these scenarios, we selected a basic reproductive number that is comparable to the values estimated for those three pandemics [24]. Second, we considered four vaccination scenarios: (*i*) a baseline of fully unmitigated transmission; (*ii*) strain-matched vaccine alone; (*iii*) Before Day One vaccine alone; and (*iv*) combined Before Day One and strain-matched vaccines. Third, we varied each of the four vaccine efficacies (Before Day One cross-protective efficacy against infection, Before Day One cross-protective efficacy against hospitalization, strain-matched efficacy against infection, strain-matched efficacy against hospitalization) over a wide range of values. Finally, we varied the time delay associated with the manufacturing/distribution of the strain-matched vaccine. Because the focus of this work is on vaccination, we did not incorporate behavior modifications or non-pharmaceutical interventions (NPIs). Additional details about the model, parameter values chosen, scenario designs, etc., are provided in the supplementary material.

## Results

A core assumption of this work is that the Before Day One vaccine provides modest protection against emerging viruses with pandemic potential. In contrast, the strain-matched vaccine is assumed to be more efficacious. For this reason, we first focus on two scenarios of Before Day One vaccine efficacy: a low vaccine effectiveness (VE) scenario, in which the vaccine efficacies against infection and hospitalization with the pandemic strain are 10% and 30%, respectively; and a moderate VE scenario, in which the corresponding values are 20% and 40%. The efficacies of the strain-matched vaccine are 50% against infection and 80% against hospitalization. The HHS Pandemic Influenza Plan targets the deployment of strain-matched vaccines to the population within 12 weeks of a pandemic declaration [1]. To account for the time necessary for strain matching and production of the pandemic vaccine, we optimistically assumed that the strain-matched vaccine begins distribution exactly 12 weeks after the first one hundred infections in the U.S., even though the pandemic declaration may not coincide with this early stage of spread. The supplementary material includes results for both earlier and later deployment of the strain-matched vaccine.

To demonstrate the impact of Before Day One as well as strain-matched vaccination on an influenza pandemic, we compare the weekly hospitalization incidence across the three pandemic scenarios and two VE scenarios (Fig. 1). The figure shows that due to its delayed availability, the strain-matched vaccine, on its own, provides no substantial mitigation in hospitalizations, unless viral transmission is low enough to produce a delayed pandemic peak (2009-like scenario) or NPIs are applied (see Supplement). In contrast, the Before Day One vaccine on its own provides protection from the onset and produces noticeable mitigation of hospitalizations at all levels of transmission considered here. Combining both vaccines does not provide additional benefits in the medium and high transmission scenarios considered (1968-like and 1918-like), since the hospitalization incidence peaks when the strain-matched vaccine coverage is still too low.

**Figure 1:**
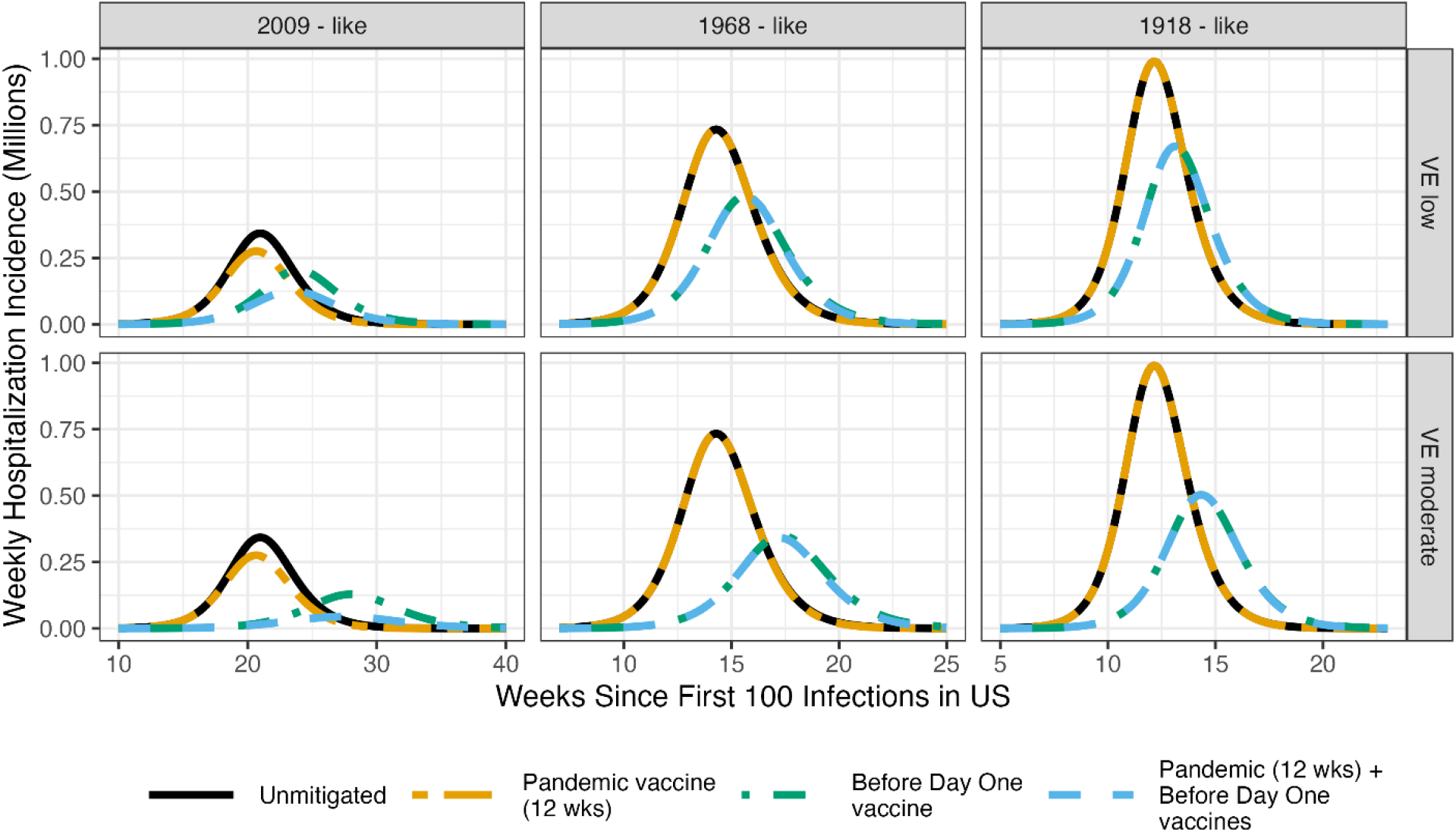
Weekly hospitalization incidence after the first 100 infections in the U.S., for different pandemic scenarios (columns) and Before Day One vaccine efficacy against the pandemic virus (rows). Each curve corresponds to a different mitigation strategy: unmitigated, strain-matched vaccine alone, seasonal Before Day One vaccine alone and both vaccines combined. Strain-matched vaccine has efficacies of 50% and 80% against infection and hospitalization, respectively. The Before Day One vaccine has a coverage of 48% over the whole population and efficacies of 10/30% (top row) and 20/40% against infection/hospitalization (bottom row). Note the difference in time span covered for each pandemic scenario. For the 1968- and 1918-like scenarios, there is near perfect overlap between: (*i*) the unmitigated and strain-matched vaccine alone scenarios and (*ii*) the Before Day One vaccine alone and strain-matched plus Before Day One vaccine scenarios.

To further quantify the impact of Before Day One vaccination, we excluded the strain-matched vaccine and examined the seasonal efficacy space in greater detail. We calculated the total number of hospitalizations (Fig. 2) over the pandemic duration as a function of the efficacies against infection and hospitalization. The Before Day One vaccine alone, at the two efficacy scenarios considered in Fig. 1, reduces the total number of hospitalizations to the values shown in Table 1. This corresponds to a reduction of about 30% in the low VE scenario and above 40% in the moderate VE scenario. The steepness of the contour lines in Fig. 2 indicates that the efficacy against infection has a higher impact in mitigating the number of hospitalizations, compared to the efficacy against hospitalization. This reflects the fact that protection against infection directly limits transmission, preventing vaccinated individuals from contributing to onward spread. In contrast, the vaccine efficacy against hospitalization only improves the clinical outcomes for the recipient.

**Table 1:** Total hospitalizations for each pandemic and intervention scenario. The low VE scenario consists solely of a Before Day One vaccine with efficacies equal to 10% against infection and 30% against hospitalization. The efficacies for the moderate VE scenario are 20% and 40%, respectively. We assume that 48% of the total population of 338M has received the Before Day One vaccine.

| ***Pandemic scenario*** | ***Unmitigated*** | ***Low VE (10/30)*** | ***Moderate VE (20/40)*** |
| --- | --- | --- | --- |
| 2009-like | 2.3M | 1.6M | 1.2M |
| 1968-like | 3.2M | 2.3M | 1.9M |
| 1918-like | 3.7M | 2.7M | 2.2M |

**Figure 2:**
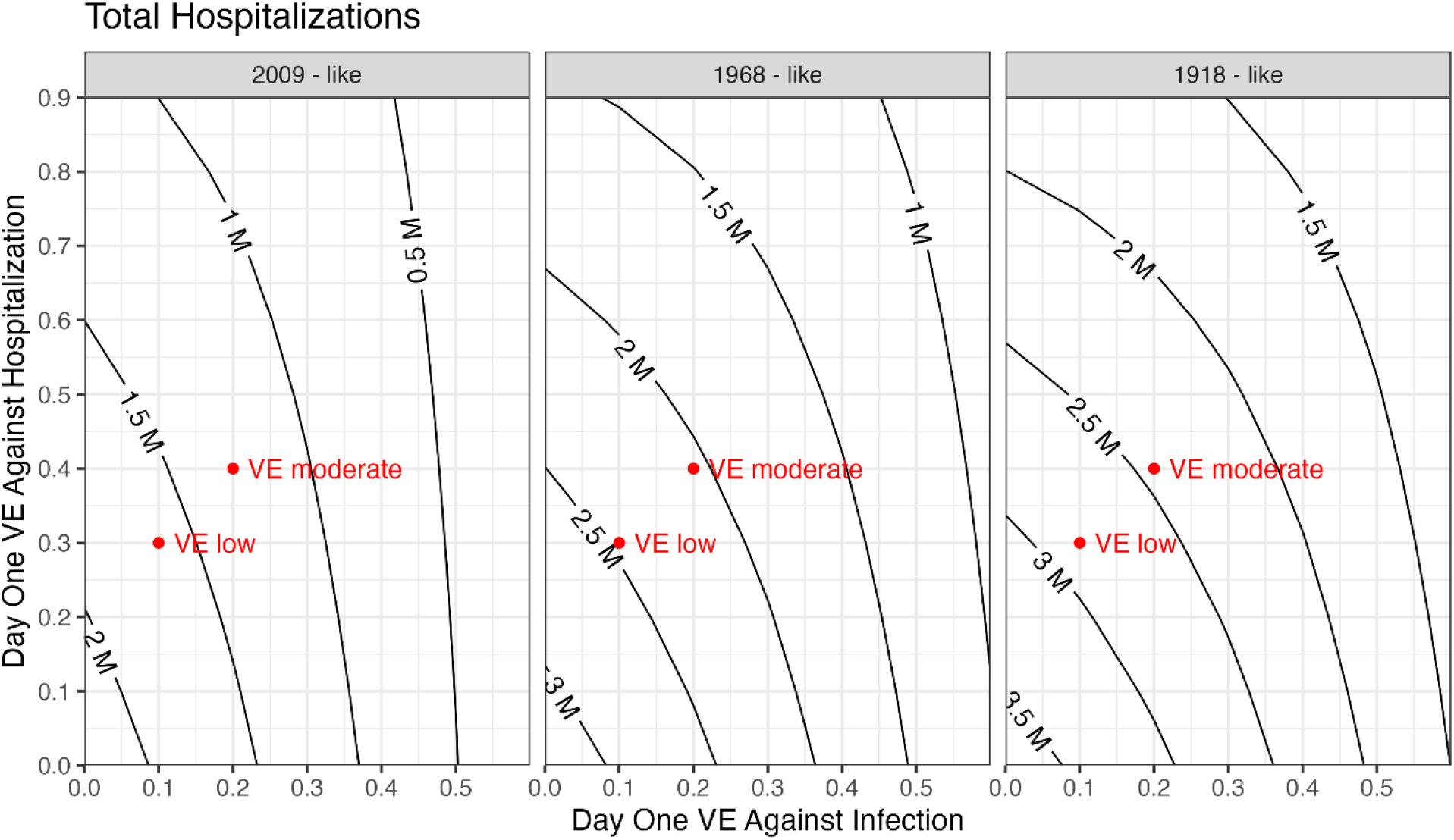
Total hospitalizations, measured in millions, for the mitigation scenario of “Before Day One vaccine alone” across different values of the efficacies against infection (x-axis) and against hospitalization (y-axis) for different levels of viral transmissibility (panels). The points in each panel mark the location of the low and moderate VE scenarios of Fig. 1.

The results presented thus far focus on the population impact of a cross-protective Before Day One vaccine. However, the decision to receive a vaccine rests, at least partially, on individual-level benefits. To assess this, we investigate how the hospitalization rate—the fraction of a group that is hospitalized at any point before the end of the pandemic —varies by age-group and vaccination status. We then calculate the percent reduction in hospitalization rate between vaccinated and unvaccinated individuals, averaged across age groups (Fig. 3). At the low and moderate VE scenarios shown in Fig. 1, vaccinated individuals experience hospitalization rates that are 35-50% lower than those of their unvaccinated counterparts.

**Figure 3.**
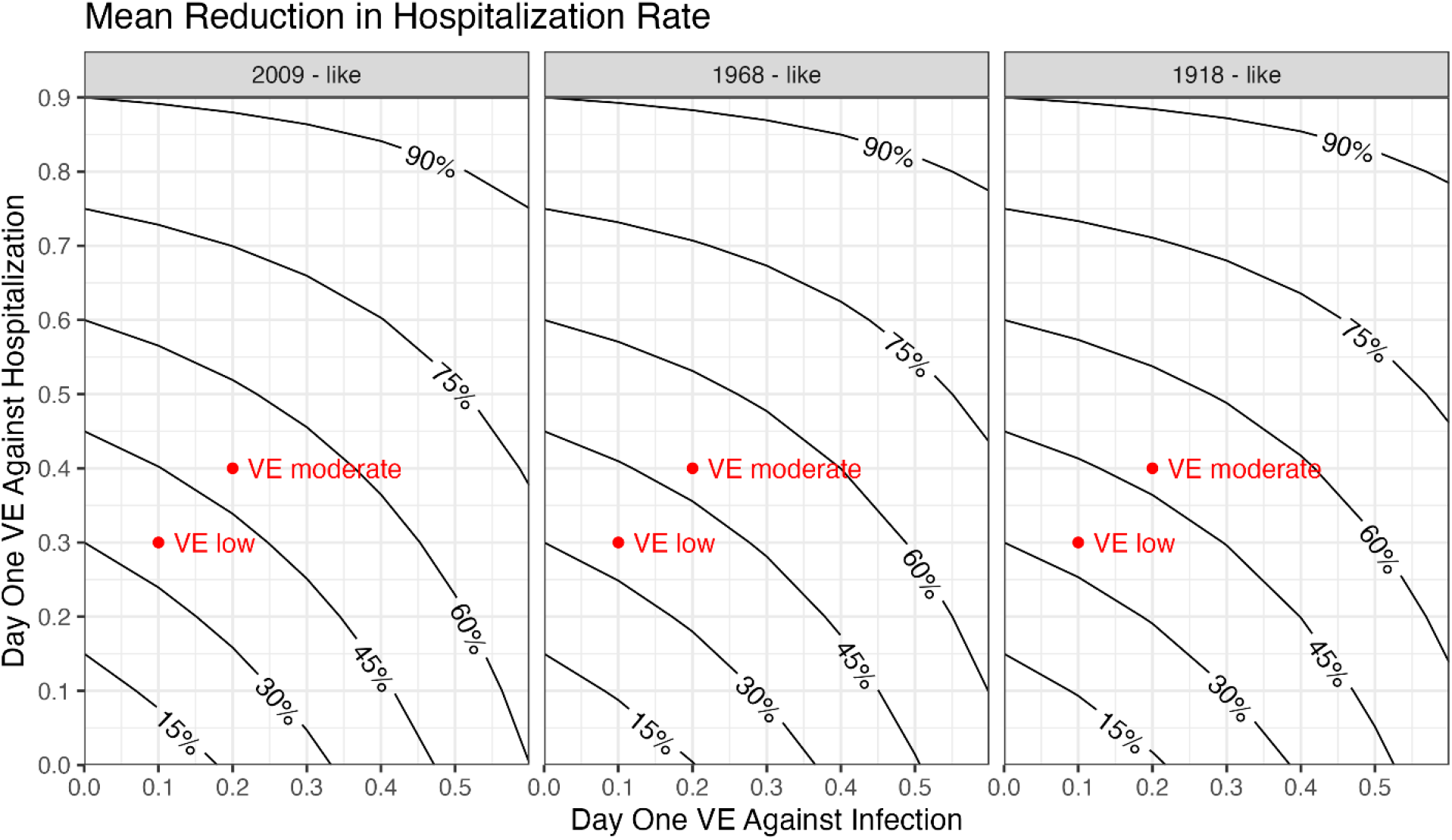
Percent reduction in hospitalization rate between vaccinated and unvaccinated individuals, averaged across age groups, for the scenario of “Before Day One vaccine alone”. The axes correspond to the efficacies against infection (x-axis) and against hospitalization (y-axis) for different levels of viral transmissibility (panels). The points in each panel mark the location of the low and moderate VE scenarios of Fig. 1.

## Discussion

We evaluated the impact of a “Before Day One vaccine” – a seasonal influenza vaccine at typical uptake levels that provides broad, but modest efficacy, against emerging influenza viruses with pandemic potential. We compared this mitigation strategy with the more standard one of relying on a pandemic strain-matched vaccine. Our results show that even under low and moderate Before Day One vaccine pandemic efficacy assumptions, seasonal Before Day One vaccination can provide the significant population-level benefit of decreasing hospitalization by more than 30%. At the individual level, the hospitalization rate among recipients goes down by 35-50% when compared to the unvaccinated.

Pandemic strain-matched vaccines can be highly efficacious against the virus variant in circulation. In contrast to the situation with single-peak epidemics studied here, a matched vaccine can be highly impactful during later stages of a protracted or multi-peaked epidemic. However, the extended time required for the development, manufacturing, and distribution of matched vaccines greatly limits their early impact soon after viral emergence. For example, during the 2009 pandemic, the first cases in the US were detected in April and a public health emergency was declared shortly thereafter, yet mass vaccination did not begin until October [25]. For this reason, it is necessary to develop mitigation tools that are tailored to have an impact from day one of pandemic virus emergence, such as the one investigated here.

For the results shown here, we made the optimistic choice of distributing the strain-matched vaccine at 12 weeks after the first one hundred infections in the U.S. With this distribution delay, the strain-matched vaccine is only able to mitigate hospitalizations at the lowest level of viral transmission explored here (2009-like). In the supplement, we explore scenarios with strain-matched vaccine distribution delays that are either shorter or longer than 12 weeks (Fig. S5). These results show two things: (*i*) that extremely rapid manufacturing and distribution are necessary to even minimally mitigate the 1968- and 1918-like scenarios and (*ii*) that distribution delays longer than 12 weeks cause the strain-matched vaccine to provide minimal benefit at any transmission level explored in this work. A second assumption that impacts our results is the assumed durability of the Before Day One vaccine’s protection. Since we have not incorporated efficacy waning into our modeling, the tacit assumption is either (*i*) that the duration of protection against pandemic viruses is longer than a seasonal vaccination cycle or (*ii*) that the efficacies do wane over the duration of one season but reach the effective values studied here by the time the pandemic virus emerges (i.e., 10/30% and 20/40% in the low and moderate VE scenarios, respectively). We leave a more thorough exploration of Before Day One vaccine waning to future investigations.

We also assume that the seasonal Before Day One vaccine coverage is at seasonal maximums at the time of virus emergence. Our results would be affected should a pandemic virus emerge in the early or middle phases of seasonal influenza vaccination campaign, when coverage has not yet plateaued and if Before Day One vaccine protection were of sub-yearly durability. In this situation, protection against the emerging pandemic virus would arise partly from the current and prior seasonal vaccine.

There are several elements that we did not include in our primary analysis. First, in order to maintain a narrow focus on vaccine impact, we did not consider non-pharmaceutical interventions (NPIs) nor spontaneous behavior changes that might emerge in response to an emerging influenza pandemic. Any behavior change that might occur to reduce transmissibility would have the effect of slowing transmission down or, more colloquially, flattening the curve [26, 27, 28, 29, 30, 31]. In the supplement, we include scenarios in which implemented NPIs consistent with those observed during COVID-19 [30, 31] result in a delayed peak and sufficient time for the matched vaccine to be broadly distributed and make a meaningful impact (Fig. S6).

Another limitation is that our model does not account for the effects of recent seasonal influenza infections or vaccination on the complexity of the immune landscape at the beginning of a pandemic. For example, in the early months of the 2009 pandemic, epidemiological studies from Canada reported the paradoxical finding that vaccination with the seasonal trivalent vaccine increased the odds of symptomatic illness with the AH1N1pdm09 virus [32]. A proposed explanation was that individuals without the trivalent vaccine were more likely to undergo natural infection with the seasonal variant and that this provided them with a broader cross-protection than uninfected trivalent vaccine recipients [33]. Other modeling efforts have incorporated some aspects of the complex immune interplay between multiple circulating strains and the vaccines intended to protect against them [34, 35, 36, 37]. However, we deliberately tailored our model to focus solely on the suppression of the pandemic virus’ transmission.

In summary, our results provide evidence that a seasonal Before Day One vaccine with broad cross-reactivity would have a substantial impact on pandemic mitigation, even at modest vaccine efficacies and typical seasonal vaccine uptake rates. However, since Before Day One vaccines are still under development and many approaches use new technology, there is as of now no data to inform what the range of possible vaccine efficacies would be, especially across the spectrum of possible emerging variants. In addition, since it is not possible to carry out vaccine trials against variants that have yet to emerge, the efficacies against such hypothetical variants would need to be inferred from animal models and other immunogenicity data. Despite these challenges, our results suggest that broadly protective seasonal Before Day One vaccines hold the promise of becoming a key tool for U.S. pandemic influenza preparedness.

## Data Availability

All data produced in the present work are contained in the manuscript

