## supplement for "Pandemic Influenza Protection Before Day One: Modeling the Impact of a Broadly Cross-Reactive Seasonal Vaccine"

**Supplementary Material**

**Study Population**

Our model assumes the demographic structure of the US in 2024 (total: 339million), using the grouping by age: 0-4 yrs, 5-17 yrs, 18-49 yrs, 50-64 yrs and 65+ yrs. The population data is obtained from the American Community Survey (ACS) [1], using the R library tidycensus [2].

**Parameter Assumptions**

For results presented in the main text, we assume epidemiological parameter values as listed in Table S1, whereas vaccine parameters used are listed in Table S2. Note that we assume no efficacy waning over the course of a single season/epidemic wave. Parameters that vary by age group are listed in Table S3. The seasonal vaccine coverage by age is assumed to reflect that of recent influenza seasons [3, 4].

| **Parameter** | **Value/Assumptions** | **Reference** |
| --- | --- | --- |
| Latent period | Exponentially distributed with a mean of 2.5 days | [5, 6, 7] |
| Infectious period | Exponentially distributed with a mean of 1.5 days | [5, 6, 7] |
| Basic reproductive number | Three pandemic scenarios considered with basic reproductive numbers: 1.8 (1918-like), 1.64 (1968-like) and 1.4 (2009-like). | [8] |
| Probability of infectious contact from source to target age | Determined by contact matrix | [9] |

**Table S1:** epidemiological parameters used in the model.

**Flumodels**

Flumodels is an R library for modeling infectious disease transmission [10]. For the current work, we consider a susceptible, exposed, infectious, and recovered compartmental model (SEIR). The compartments are further subdivided to account for the distribution of a vaccine (Fig. S1).

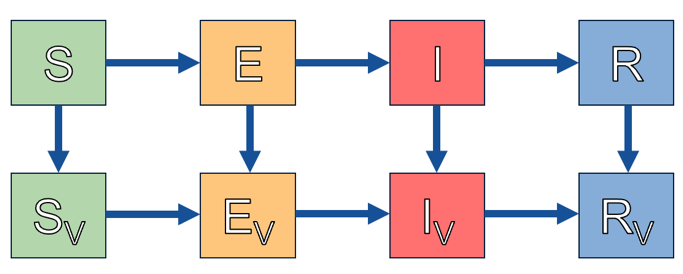

**Figure S1:** Flow diagram for compartmental model, with the compartments: susceptible (S), exposed (E), infected (I), and recovered (R). Compartments with a ‘V’ subscript denote vaccination. Each compartment is further divided into age groups (not shown).

| **Vaccine parameter** | **Value/Assumptions** | **Reference** |
| --- | --- | --- |
| Seasonal vaccine efficacy against infection | 0-60% | – |
| Seasonal vaccine efficacy against hospitalization | 0-90%, in steps of 10% | – |
| Pandemic-matched vaccine efficacy against infection | 50% | [11] |
| Pandemic-matched vaccine efficacy against hospitalization, conditional on breakthrough infection | 80% | [12, 13, 14, 15, 16] |
| Delay for pandemic-matched vaccine to become efficacious | 4 weeks | [17, 18] |
| Pandemic-matched vaccine doses applied per day | 2 million | [19] |

**Table S2:** vaccination-related parameters used in the model.

**Hospitalizations**

It is assumed that out of all unvaccinated infections, an age-dependent fraction given by the Infection Hospitalization Rate (IHR) and with values given in Table S3, will ultimately require hospitalization. We do not account for time delays between infection and hospitalization, nor do we keep track of hospitalization duration. The fraction of vaccine recipients who require hospitalization is covered in the next two sections.

**Pandemic-matched and Seasonal Vaccines Model**

*Pandemic matched vaccination*

In this model, we only explicitly track doses of the pandemic-matched vaccine (Fig S2). Although the model only considers a single dose of this vaccine, we mimic the action of a two-dose vaccine by having a delay of 4 weeks before efficacy takes effect: 3 weeks between doses and 1 more for efficacy ramp up. Conceptually, our model thus tracks full courses rather than individual doses. The timing chosen reflects the speed of an immunological response after vaccination and the schedule between doses during the early phases of the COVID19 pandemic [17, 18].

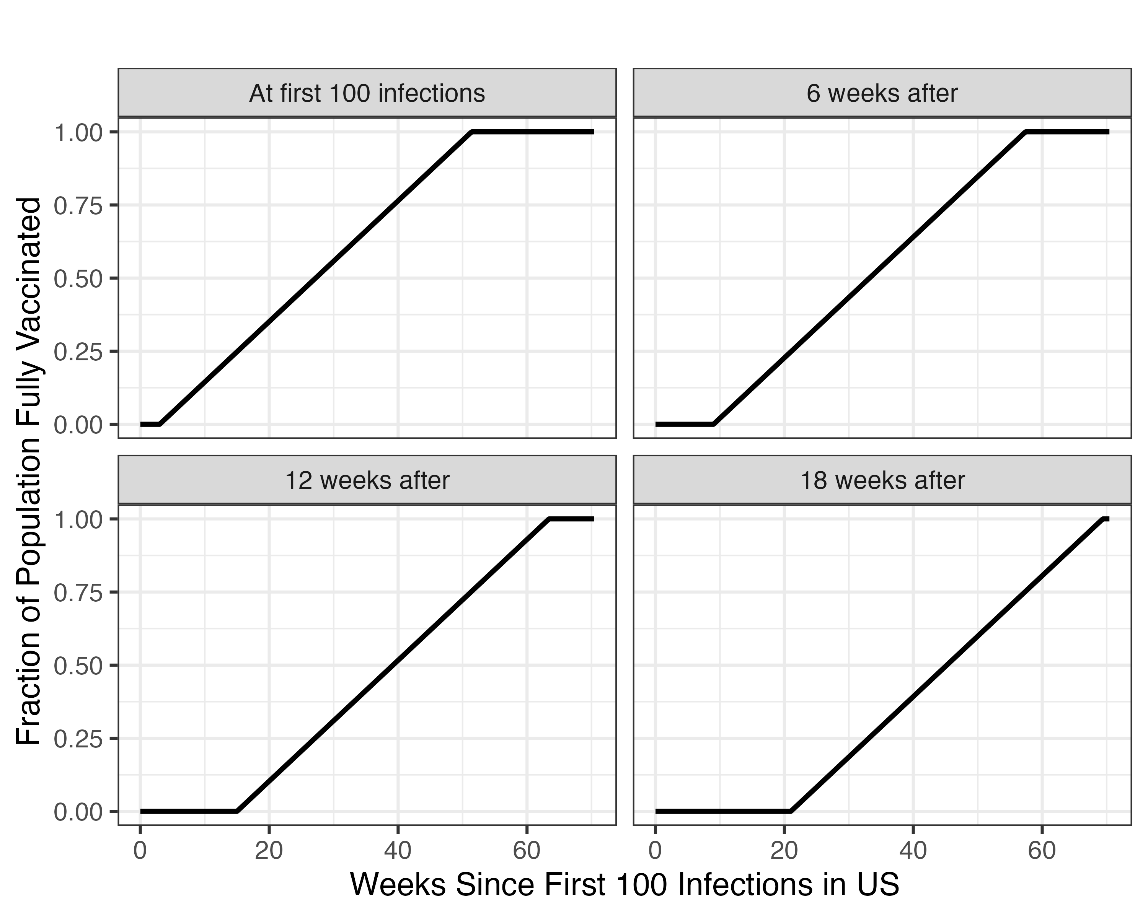

**Figure S2:** Distribution of pandemic-matched vaccine to the population when release happens at different timepoints: coinciding with the first one hundred infections and every 6 weeks after that up to 18 weeks. A 3-week delay between doses is assumed. At a rate of 1 million courses per day, it takes roughly 48 weeks to achieve full population coverage.

Daily vaccine doses administered during the COVID-19 pandemic show high variability in time [19], with a 1 week rolling mean maximum of 3.3 million doses attained in a single day in April of 2021 (Fig S3). The rate of administration depends upon many factors including vaccine availability, administration capacity, vaccination willingness, etc. Note that the highest vaccination rate during the COVID-19 pandemic occurred soon after the vaccines obtained Emergency Use Authorizations from the FDA in December of 2020 and February of 2021. Between February and June of 2021, more than 1 million vaccine doses were administered per day, with an average of 2 million, roughly. Thus, we assume that once the pandemic-matched vaccine becomes ready for mass-distribution, there is sufficient availability and capacity to apply it to the population at a rate of 2 million injections per day, or 1 million courses per day, per our model’s reckoning.

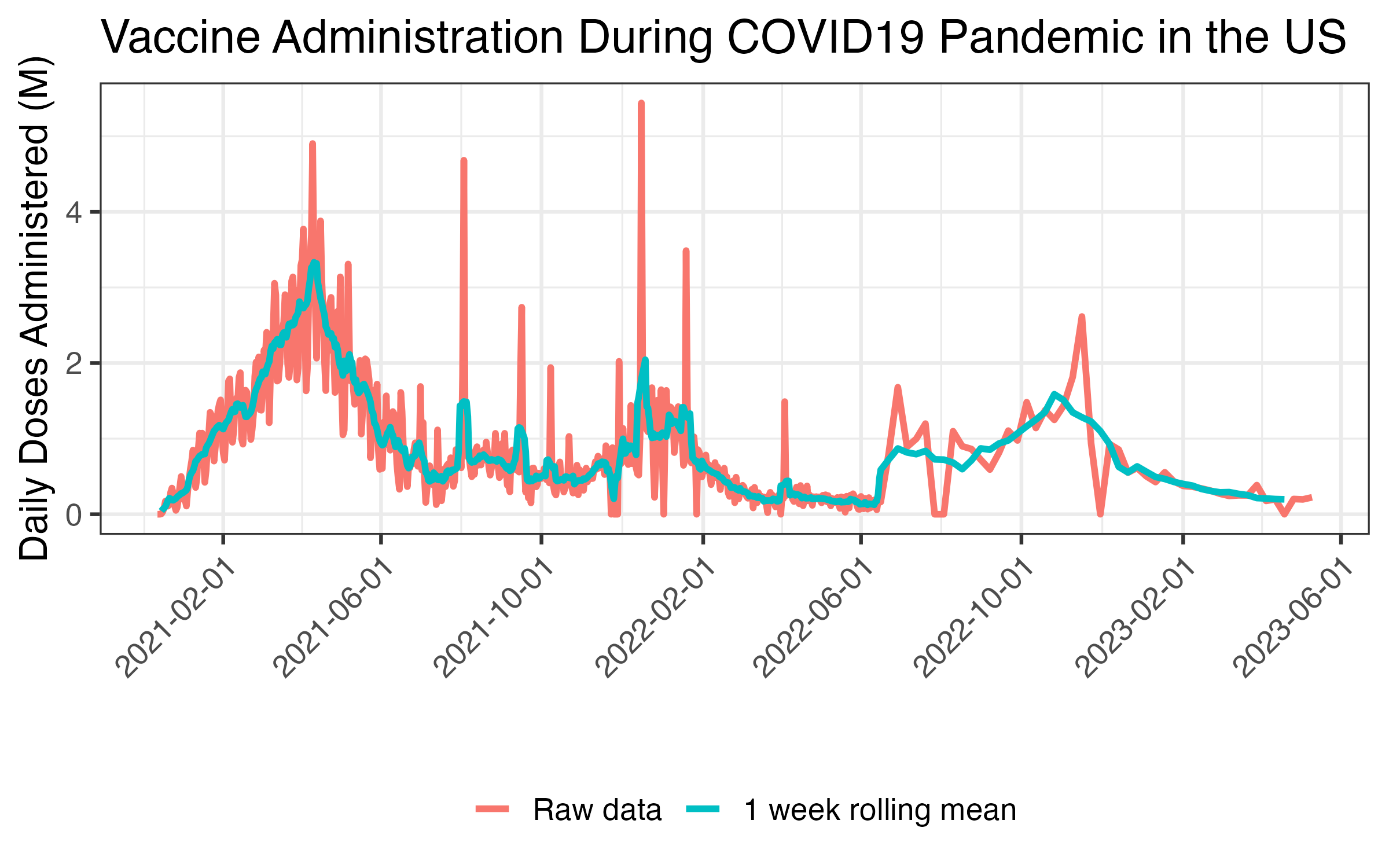

**Figure S3:** Daily vaccine doses administered during the COVID-19 pandemic in the U.S. [19].

| **Age Range** | **Number of individuals in millions**  **(% of total)** | **Number of hospitalizations per 1000 unvaccinated infections** | **Seasonal Vaccine Coverage** |
| --- | --- | --- | --- |
| 0 – 4yrs | 18.92M (5.58%) | 5 | 60% |
| 5 – 17yrs | 55.25M (16.30%) | 3 | 49% |
| 18 – 49yrs | 142.31M (42.00%) | 8 | 36% |
| 50 – 64yrs | 63.88M (18.85%) | 19 | 49% |
| 65+ yrs | 58.48M (17.26%) | 84 | 69% |

**Table S3**: Demographic data from [1]; hospitalization rates from [20, 21, 22, 23, 24, 25, 26]; and seasonal vaccine coverage from [3, 4].

*Seasonal vaccination*

The seasonal vaccine is distributed across age groups assuming the coverage given in Table S3. For those who receive the seasonal vaccine, the impact is twofold:

1. To model seasonal cross-protection against infection by a pandemic strain, we initiate the model – for each age group a – with a fraction $V_{a}\cdot{VE}_{S}^{seasonal}$ of susceptibles moved to the recovered class, indicating successful immunizing vaccination. Here, $V_{a}$,denotes the vaccine uptake for age group $a$ This represents the portion of vaccine recipients who, because of seasonal vaccine action, become immune to infection. A fraction ${1- V}_{a}\cdot{VE}_{S}^{seasonal}$ remain in the susceptible compartment representing the unvaccinated (a fraction ${1-V}_{a}$) plus those vaccinated for whom the vaccine fails to provide full infection immunity (a fraction $V_{a}\cdot{(1-VE}_{S}^{seasonal})$. See Fig. S4.
2. To model seasonal cross-protection against hospitalization among breakthrough infections, we first let ${IHR}_{a}$ denote the Infection Hospitalization Rate for age group $a$. For age group $a$, a fraction ${1- V}_{a}\cdot{VE}_{S}^{seasonal}$ will initially be susceptible either because they are unvaccinated or because the seasonal vaccine was not effective in providing infection immunity (Fig. S4). Among these susceptibles, a fraction $V_{a}\cdot{(1-VE}_{S}^{seasonal})$ received a vaccine that failed to protect against infection. Thus, a fraction $f_{a}$ of the susceptibles at age a will have received the seasonal vaccine but not been conferred immunity from infection, with:

$f_{a}= \frac{V_{a}\cdot(1-{VE}_{S}^{seasonal})}{{1- V}_{a}\cdot{VE}_{S}^{seasonal}}$.

Correspondingly, a fraction $f_{a}\cdot{VE}_{H}^{seasonal}$, will be conferred protection against hospitalization and the complement, $1-f_{a}\cdot{VE}_{H}^{seasonal}$, will not. Therefore, among those who do not receive the pandemic-matched vaccine and eventually become infected, a fraction:

$IHR_{a}\cdot\left( 1-f_{a}\cdot{VE}_{H}^{seasonal} \right)$*,*

go on to need hospitalization. In contrast, the recipients of both vaccines have an additional layer of protection and of them, the ones who become infected have a probability:

$$IHR_{a}\cdot\left( 1-{VE}_{H}^{pandemic} \right)\cdot\left( 1-f_{a}\cdot{VE}_{H}^{seasonal} \right)$$

of eventually becoming hospitalized.

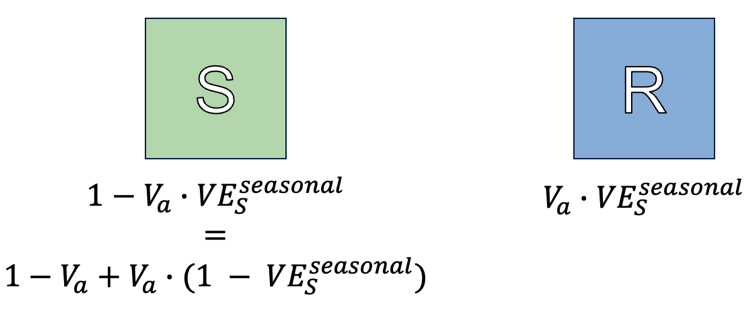

**Figure S4:**. Breakup of the age group fraction that is initially in the susceptible and recovered compartments, respectively, due to the sterilizing action of the seasonal vaccine

**Purely Seasonal Vaccine Model**

In this model, we explicitly track the doses of seasonal vaccine distributed in the population. To reflect the seasonal nature of this vaccine, it is applied up to the coverage indicated in Table S3 and much before infections get seeded into the population. Among those who do not receive the seasonal vaccine, a fraction ${IHR}_{a}$ of infections, varying by age, will require hospitalization. In contrast, for those who do receive the seasonal vaccine, a fraction

$$IHR_{a}\cdot\left( 1-{VE}_{H}^{seasonal} \right)$$

of the infected will ultimately become hospitalized.

**Exploration of Pandemic Vaccine Scenarios**

In this section, we explore the pandemic vaccine parameter space in more detail. We calculate the total hospitalizations during the course of the pandemic for different parameters of the pandemic vaccine. For mitigation, we only consider an unmitigated scenario and a pandemic vaccine alone scenario. We vary the vaccine distribution delay and the efficacies against infection and hospitalization, for the three pandemic scenarios considered (Fig. S5).

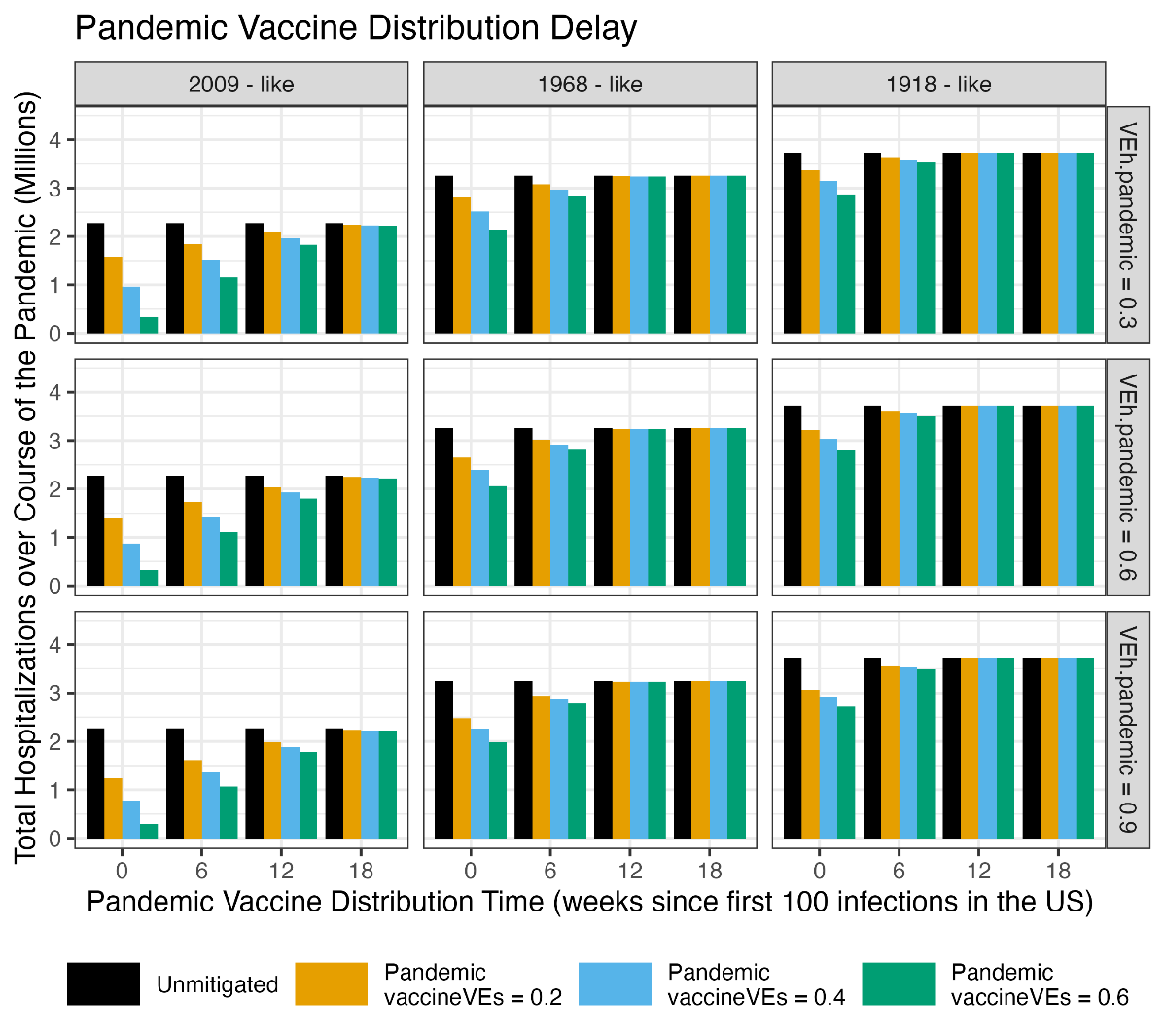

**Figure S5:** Total hospitalizations over the course of the pandemic as a function of the pandemic vaccine distribution delay, relative to the occurrence of the first 100 infections in the U.S. We consider different pandemic scenarios (columns), different pandemic vaccine efficacies against hospitalization (rows), and different mitigation strategies and efficacies against infection (colored curves/symbols).

**Delay of the Pandemic Peak via Non-Pharmaceutical Interventions**

Here, we explore how the implementation of Non-Pharmaceutical Interventions (NPIs) can be utilized to delay the pandemic peak sufficiently for the matched vaccine to have a substantial mitigating impact. For demonstration purposes, we consider an NPI implemented uniformly during a span of 12 weeks and that results in a reduction of the infectious contact rate of 30% across all age groups (Fig. S6), consistent with the evaluated impact of social distancing during the COVID-19 pandemic [27, 28]. As we see from the results, an NPI of this nature delays the peak and allows the matched vaccine to have a substantial impact if released by week 12 or earlier, for all three pandemic scenarios considered. However, more aggressive NPIs (or other mitigating elements) would be needed for the matched vaccine to have an impact if released on week 18 after the first hundred infections.

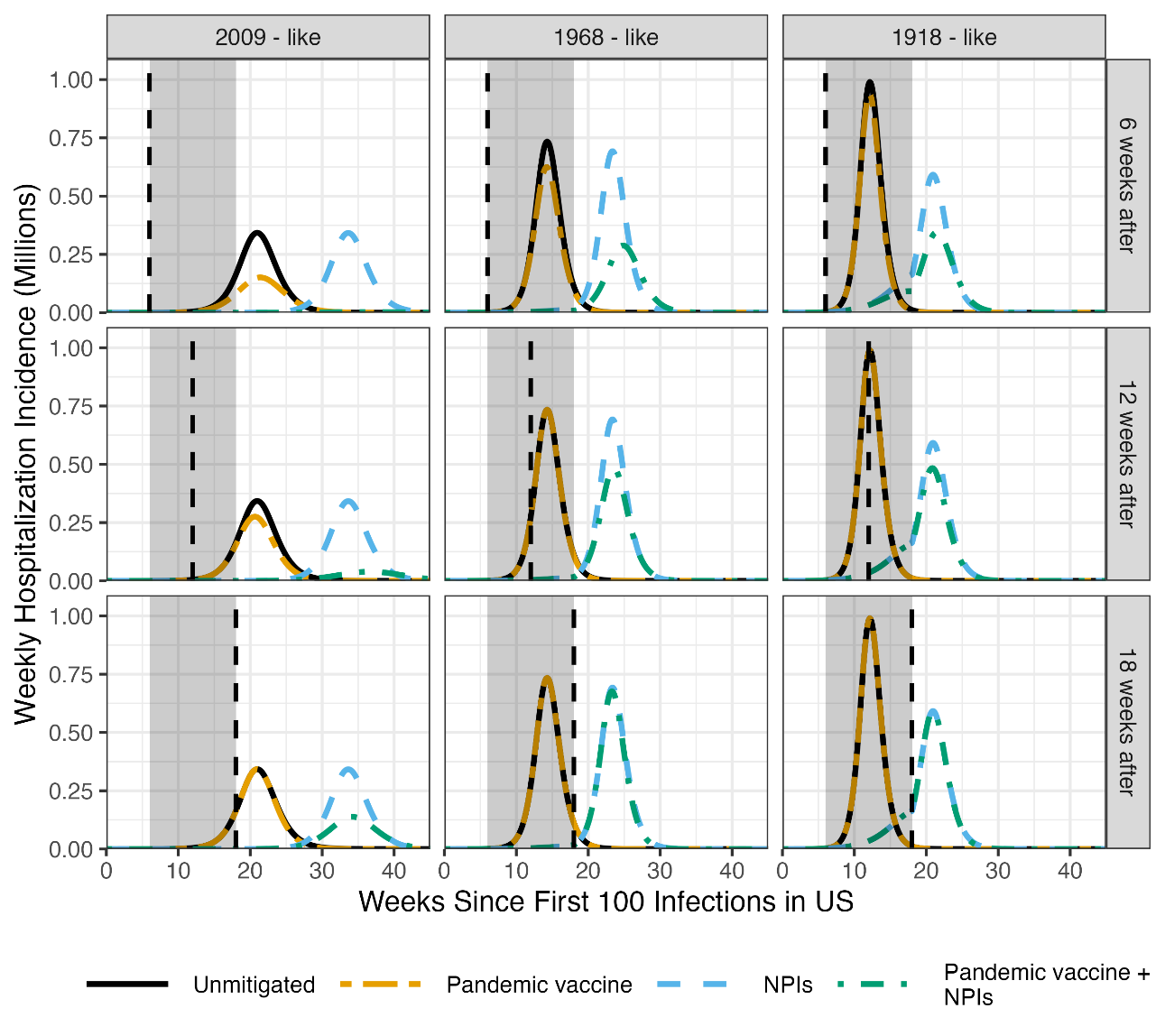

**Figure S6:** Weekly hospitalization incidence after the first 100 infections in the U.S., for different pandemic scenarios (columns) and starting week of the matched-vaccine distribution, shown with the vertical dashed line (rows). Each curve corresponds to a different mitigation strategy: unmitigated, strain-matched vaccine alone, NPIs alone, and NPIs together with matched vaccine. Strain-matched vaccine has efficacies of 50% and 80% against infection and hospitalization, respectively. The “NPIs” (dashed blue) and the “Pandemic vaccine + NPIs” (dash-dotted green) scenarios are subject to a 30% reduction in infectious contact rate from the baseline between weeks 6 and 18 (shaded area), for all age groups.

### **Bibliography**

| [1] | United States Census Bureau, "American Community Survey (ACS)," 2024. [Online]. Available: https://www.census.gov/programs-surveys/acs.html. |
| --- | --- |
| [2] | K. Walker and H. Matt, *tidycensus: Load US Census Boundary and Attribute Data as 'tidyverse' and 'sf'-Ready Data Frames,* R package version 1.7.5, 2026. |
| [3] | Centers for Disease Control and Prevention, "Weekly Cumulative Influenza Vaccination Coverage by Flu Season, Selected Demographics, and Race and Ethnicity Among Children 6 Months-17 Years, United States," March 2026. [Online]. Available: https://data.cdc.gov/Child-Vaccinations/Weekly-Cumulative-Influenza-Vaccination-Coverage-b/vncy-2ds7/about_data. |
| [4] | Centers for Disease Control and Prevention, "Weekly Influenza Vaccination Coverage and Intent for Vaccination, Overall, by Selected Demographics and Jurisdiction, Among Adults 18 Years and Older," March 2026. [Online]. Available: https://data.cdc.gov/Flu-Vaccinations/Weekly-Influenza-Vaccination-Coverage-and-Intent-f/sw5n-wg2p/about_data. |
| [5] | A. Cori, A. Valleron, F. Carrat, T. Scalia, G. Thomas and P. Boëlle, "Estimating influenza latency and infectious period durations using viral excretion data," *Epidemics,* vol. 4, no. 3, pp. 132-8, 2012. |
| [6] | M. Saito, N. Hirotsu, H. Hamada, M. Takei, K. Honda, T. Baba, T. Hasegawa and Y. Kitanishi, "Reconstructing the household transmission of influenza in the suburbs of Tokyo based on clinical cases," *Theor Biol Med Model,* vol. 18, no. 1, 2021. |
| [7] | P. Boëlle, S. Ansart, A. Cori and A. Valleron, "Transmission parameters of the A/H1N1 (2009) influenza virus pandemic: a review," *Influenza Other Respir Viruses,* vol. 5, no. 5, pp. 306-16, 2011. |
| [8] | M. Biggerstaff, S. Cauchemez and C. Reed, "Estimates of the reproduction number for seasonal, pandemic, and zoonotic influenza: a systematic review of the literature," *BMC Infect Dis,* vol. 14, no. 480, 2014. |
| [9] | J. Mossong, N. Hens, M. Jit, P. Beutels, K. Auranen, R. Mikolajczyk, M. Massari, S. Salmaso, G. Scalia, J. Wallinga, J. Heijne, M. Sadkowska-Todys, M. Rosinska and J. John Edmunds, "Social Contacts and Mixing Patterns Relevant to the Spread of Infectious Diseases," *PLoS Medicine,* vol. 5, no. 3, 2008. |
| [10] | Medical-Countermeasures Quantitative Analysis Team, "Flumodels," 2017. [Online]. Available: https://github.com/HHS/ASPR-flumodels/tree/master. |
| [11] | N. Basta, M. Halloran, L. Matrajt and I. Longini, "Estimating influenza vaccine efficacy from challenge and community-based study data," *Am J Epidemiol.,* vol. 168, no. 12, pp. 1343-52, 2008. |
| [12] | J. Puig-Barberà, A. Arnedo-Pena, F. Pardo-Serrano, M. Tirado-Balaguer, S. Pérez-Vilar, E. Silvestre-Silvestre, C. Calvo-Mas, L. Safont-Adsuara and M. Ruiz-García, "Effectiveness of seasonal 2008-2009, 2009-2010 and pandemic vaccines, to prevent influenza hospitalizations during the autumn 2009 influenza pandemic wave in Castellón, Spain. A test-negative, hospital-based, case-control study," *Vaccine,* vol. 28, no. 47, pp. 7460-7, 2010. |
| [13] | E. Belongia, M. Simpson, J. King, M. Sundaram, N. Kelley, M. Osterholm and H. McLean, "Variable influenza vaccine effectiveness by subtype: a systematic review and meta-analysis of test-negative design studies," *Lancet Infect Dis.,* vol. 16, no. 8, pp. 942-51, 2016. |
| [14] | P. Hardelid, D. Fleming, J. McMenamin, N. Andrews, C. Robertson, P. SebastianPillai, J. Ellis, W. Carman, T. Wreghitt, J. Watson and R. Pebody, "Effectiveness of pandemic and seasonal influenza vaccine in preventing pandemic influenza A(H1N1)2009 infection in England and Scotland 2009-2010," *Euro Surveill.,* vol. 16, no. 2, 2011. |
| [15] | L. Lansbury, S. Smith, W. Beyer, E. Karamehic, E. Pasic-Juhas, H. Sikira, A. Mateus, H. Oshitani, H. Zhao, C. Beck and J. Nguyen-Van-Tam, "Effectiveness of 2009 pandemic influenza A(H1N1) vaccines: A systematic review and meta-analysis," *Vaccine,* vol. 35, no. 16, pp. 1996-2006, 2017. |
| [16] | Å. Örtqvist, R. Bennet, J. Hamrin, M. Rinder, H. Lindblad, J. Öhd and M. Eriksson, "Long term effectiveness of adjuvanted influenza A(H1N1)pdm09 vaccine in children," *Vaccine,* vol. 33, no. 22, pp. 2558-61, 2015. |
| [17] | L. Montgomery and A. Larbi, "Monitoring Immune Responses to Vaccination: A Focus on Single-Cell Analysis and Associated Challenges," *Vaccines,* vol. 13, no. 4, 2025. |
| [18] | Centers for Disease Control and Prevention, "Archived COVID-19 Vaccination Schedules," Sep 2022. [Online]. Available: https://archive.cdc.gov/www_cdc_gov/vaccines/covid-19/clinical-considerations/archived-covid-19-vacc-schedule.html. |
| [19] | Centers for Disease Control and Prevention, "COVID-19 Vaccinations in the United States,County," 2023. [Online]. Available: https://data.cdc.gov/Vaccinations/COVID-19-Vaccinations-in-the-United-States-County/8xkx-amqh/about_data. |
| [20] | J. Wu, A. Ho, E. Ma, C. Lee, D. Chu, P. Ho, I. Hung, L. Ho, C. Lin, T. Tsang, S. Lo, Y. Lau, G. Leung, B. Cowling and J. Malik, "Estimating Infection Attack Rates and Severity in Real Time during an Influenza Pandemic: Analysis of Serial Cross-Sectional Serologic Surveillance Data," *PLoS Medicine,* vol. 8, no. 10, 2011. |
| [21] | J. Wu, E. Ma, C. Lee, D. Chu, P. Ho, A. Shen, A. Ho, I. Hung, S. Riley, L. Ho, C. Lin, T. Tsang, S. Lo, Y. Lau, G. Leung, B. Cowling and J. Malik, "The Infection Attack Rate and Severity of 2009 Pandemic H1N1 Influenza in Hong Kong," *Clinical Infectious Diseases,* vol. 51, no. 10, pp. 1184-1191, 2010. |
| [22] | X. Wang, C. Wong, K. Chan, K. Chan, P. Cao, J. Peiris and L. Yang, "Hospitalization risk of the 2009 H1N1 pandemic cases in Hong Kong," *BMC Infect Dis.,* vol. 14, no. 32, 2014. |
| [23] | Y. Deng, Y. Kim, A. Bratcher, J. Jones, M. Simuzingili, A. Gundlapalli, M. Hagen, R. Iachan and K. Clarke, "Ratio of Infections to COVID-19 Cases and Hospitalizations in the United States based on SARS-CoV-2 Seroprevalence Data, September 2021-February 2022," *Open Forum Infect Dis.,* vol. 12, no. 1, 2025. |
| [24] | S. Mahajan, C. Caraballo, S. Li, Y. Dong, L. Chen, S. Huston, R. Srinivasan, C. Redlich, A. Ko, J. Faust, H. Forman and H. Krumholz, "SARS-CoV-2 Infection Hospitalization Rate and Infection Fatality Rate Among the Non-Congregate Population in Connecticut," *Am J Med.,* vol. 134, no. 6, pp. 812-816, 2021. |
| [25] | N. Leung, C. Xu, D. Ip and B. Cowling, "The Fraction of Influenza Virus Infections That Are Asymptomatic: A Systematic Review and Meta-analysis," *Epidemiology,* vol. 26, no. 6, pp. 862-72, 2015. |
| [26] | Centers for Disease Control and Protection, "Flu Disease Burden: Past Seasons," 2025. [Online]. Available: https://www.cdc.gov/flu-burden/php/data-vis/past-seasons.html. |
| [27] | G. Wellenius, S. Vispute, V. Espinosa, A. Fabrikant, T. Tsai, J. Hennessy, A. Dai, B. Williams, K. Gadepalli, A. Boulanger, A. Pearce, C. Kamath, A. Schlosberg, C. Bendebury, C. Mandayam, C. Stanton, S. Bavadekar, C. Pluntke, D. Desfontaines and Jacobs, "Impacts of social distancing policies on mobility and COVID-19 case growth in the US," *Nature Commun,* vol. 12, p. 3118, 2021. |
| [28] | A. Olney, J. Smith, S. Sen, F. Thomas and H. Unwin, "Estimating the Effect of Social Distancing Interventions on COVID-19 in the United States," *Am J Epidemiol,* vol. 190, no. 8, pp. 1504-1509, 2021. |
